# Is academic medicine prepared to teach about the intersection of childhood experiences and health across the life course? An exploratory survey of faculty

**DOI:** 10.1101/2021.09.02.21263054

**Authors:** Angie Koo, Jordyn Irwin, Morgan Sturgis, Alan Schwartz, Memoona Hasnain, Elizabeth Davis, Audrey Stillerman

## Abstract

**Purpose:** Childhood experiences affect health across the lifespan. Evidence-based prevention and treatment strategies targeting early-life stress are emerging. Nevertheless, faculty physicians’ preparation to incorporate this science into practice has not been well studied. The purpose of this study was to explore medical faculty knowledge and beliefs, timing and route of knowledge acquisition, perceived relevance and application of topics, and any associated faculty characteristics.

**Method:** The research team developed and administered a 39-question survey to faculty from six departments at the University of Illinois College of Medicine and Rush Medical College in Chicago. The team employed quantitative and qualitative methods to analyze responses.

**Results:** Eighty-one (8.8%) eligible faculty completed the survey. Of respondents, 53 (65.4%) achieved a high score on knowledge questions, 34 (42.0%) on beliefs questions, and 42 (59.1%) described high concept exposure, but only six (7.4%) through a formal route. Although 78 (96.8%) respondents indicated that survey concepts are relevant, only 18 (22.2%) reported fully incorporating them in their work. Respondents reporting full incorporation of concepts were significantly more likely to attain high concept exposure scores than those not fully incorporating concepts (17 respondents, 94.4%, vs. 25 respondents, 39.7 %, P < .001), whether reporting formal or informal exposure. Both quantitative and qualitative analysis highlighted limited awareness among respondents of trauma prevalence among healthcare workers, lack of familiarity with interventions, and challenges in addressing childhood experiences given time and resource constraints.

**Conclusions:** Most respondents had some familiarity with the impact of childhood experiences on health and perceived the relevance of this science. Nonetheless, many identified the need for additional coaching. Because results suggest that exposure supports full application of concepts, intentional faculty development and establishment of medical education competencies is pivotal to prepare faculty to include these crucial topics in patient care and teaching.

---

Decades of robust basic science, public health and social science research reveal that childhood experiences, both positive and negative, shape our trajectory of health or illness across the lifespan.^1-5^ Insufficiently buffered childhood adversity and trauma, arising from experiences ranging from the individual (abuse, neglect, etc.) to structural and historical (racism, poverty, etc.), derail optimal development and impair physiologic, psychological and social function in both children and adults.^6-12^ These experiences are a common, preventable root cause of disease and disparities, including six of the ten leading causes of death in the United States (US).^13^ High rates of childhood adversity and trauma also plague health care workers.^14^ History of direct personal trauma combined with job-related vicarious and secondary trauma creates a toxic combination contributing to compassion fatigue, burnout, and suicide, with a profound impact on the healthcare workforce.^14-18^ At the same time, an emerging literature reveals that multi-layered strategies for prevention and treatment of childhood adversity and trauma include equitable distribution of resources, family support, prioritizing positive human connections, and mind-body regulatory practices.^4,19-23^

In spite of all the research, there has been a critical lag in translating this essential science into practice and integrating it into medical education, including understanding how diseases arise, creating effective strategies for prevention and treatment, and designing successful health education. As of 2016, only eight of 192 US medical schools included content on the impact of childhood trauma, which could be as little as a single lecture.^24^

Existing peer-reviewed research regarding attending physicians’ knowledge, beliefs, and practices about the impact of childhood experiences on health is limited and is almost entirely related to Adverse Childhood Experience (ACE) screening,^25-28^ which is controversial.^29-31^ Even within their narrow focus on ACEs, these studies reveal that many practicing physicians lack familiarity with the impact of childhood adversity on health. A 2013 American Academy of Pediatrics survey of 302 general pediatricians found that 76% of respondents were completely unfamiliar with the 1998 Adverse Childhood Experiences Study.^32^ A recent study of attending and resident physicians in a community with a known high prevalence of ACEs found that only 19.5% of physicians were aware of the ACE Screening Questionnaire, and only 3.5% had used it in their practice.^33^ Other studies surveyed resident physicians, nurse practitioners, and other healthcare workers, and only occasionally included attending physicians.^34-36^ Practicing healthcare workers endorsed discomfort or lack of confidence with the subject matter;^36-38^ gaps in training on ACEs and available resources,^39^ and time constraints during patient encounters as potential contributors to low screening rates.^34-36,38^

To our knowledge, no studies have yet specifically investigated academic medical faculty preparation to teach about the intersection of childhood experiences and health. To address this gap in the literature, we sought to explore the current state of 1) academic medical faculty knowledge, beliefs, timing and route of exposure to the science describing the profound impact of childhood experiences on health across the life course and emerging prevention and treatment interventions for childhood trauma; 2) perception of relevance and degree of application of these concepts to practice; 3) associated faculty characteristics; and 4) gaps requiring further training.

## Methods

### Participants and recruitment

In 2019, our interprofessional faculty and student research team selected a convenience population of 924 faculty physicians from six departments at two neighboring Chicago medical schools, the public University of Illinois College of Medicine (UIH-UIC COM) and private Rush Medical College (Rush). Criteria for participation in the study included a Doctor of Medicine (MD) or Doctor of Osteopathic Medicine (DO) degree, completion of postgraduate medical training, a faculty appointment, and listing in the UIH-UIC COM or Rush department directory for Family Medicine, Internal Medicine, Obstetrics and Gynecology, Pediatrics, Psychiatry, and/or Surgery, all of which make up the Liaison Committee on Medical Education’s required rotations in medical school.^40^ We excluded participation any faculty who contributed to study development from participation.

### Survey development and administration

In 2019 our team created an anonymous electronic survey based on key literature in epidemiology, developmental psychology, neuroscience, and traumatology.^3,5,6,11,12,14,23,41,42^ We developed, modified, and finalized the survey through three iterative cognitive interviews.^43^ See Figure 1 for the final survey which contained 39 questions: participant demographics (nine questions, multiple choice with option for multiple responses when indicated); relevant knowledge (six questions, Likert scale) and beliefs about the impact of childhood experiences on health and application of this science (11 questions, Likert scale); timing and degree of exposure to these core scientific concepts (11 questions, multiple choice with option for multiple responses); perception of concept relevance and degree of incorporation into practice (one question, multiple choice); and an optional open-ended question.

**Figure 1.** Childhood Experiences and Health Questionnaire Administered to Medical Faculty Surveyed in 2020 in Chicago, IL

In 2020, the institutional review boards of the University of Illinois at Chicago and Rush University Medical Center approved and granted exemption for this study. Eligible faculty appearing in each institution’s departmental website directory received the survey via a secure email link. The email introduced our team and project before prompting physicians to self-screen for participation via the informed consent document prior to accessing the survey. An encrypted and password protected UIC REDCap system stored all electronic data. The survey remained open over a two-week data collection period from late June through early July 2020. Faculty received email reminders one week and 48 hours before survey closure.

### Data analysis

Our team considered surveys complete if participants answered all key items. We counted participants who were faculty in two departments or at both UIC and Rush as half a participant in each for the purposes of tabulating participation rates by department or institution. We deemed a response to be “correct” or “favorable” when it represented agreement or strong agreement with a true statement or disagreement or strong disagreement with a false statement. Our team set the definition of “high” scores for knowledge and beliefs questions at ≥80% correct/favorable. We defined “high” concept exposure scores as exposure of any type (“formal”: obtained through undergraduate or medical education; “outside formal training”: obtained through journals, colleagues, conferences, popular media, family and friends or observing and listening to patients; or “exposed but can’t remember when/where I learned it”) as ≥80% of the concepts.

We summarized demographic data using descriptive statistics and performed comparisons of mean responses by groups for Likert-scaled items using t-tests and comparisons of categorical responses by groups using χ^2^ tests. We conducted analyses using R 3.6 (R Core Team, Vienna, Austria). Two authors independently reviewed, categorized and summarized themes in responses to the open-ended question using content methodology.^44^ We resolved any differences through discussion.

## Results

136 (14.7%) of 924 eligible faculty at UIH-UIC COM and Rush began this survey. We analyzed responses from the 81 (8.8%) of faculty who completed the survey. Response rate was 44 of 257 (17.7%) eligible respondents at UIC and 36 of 667 (5.5%) at Rush (P < .001). Response rate also varied by department; Pediatrics had the highest rate (31 of 123 eligible respondents, 24.0%), followed by Family Medicine (12 of 63 eligible respondents, 18.3%), and Internal Medicine (33 of 405 eligible respondents, 7.4%). See Table 1 for respondent characteristics.

**Table 1.**
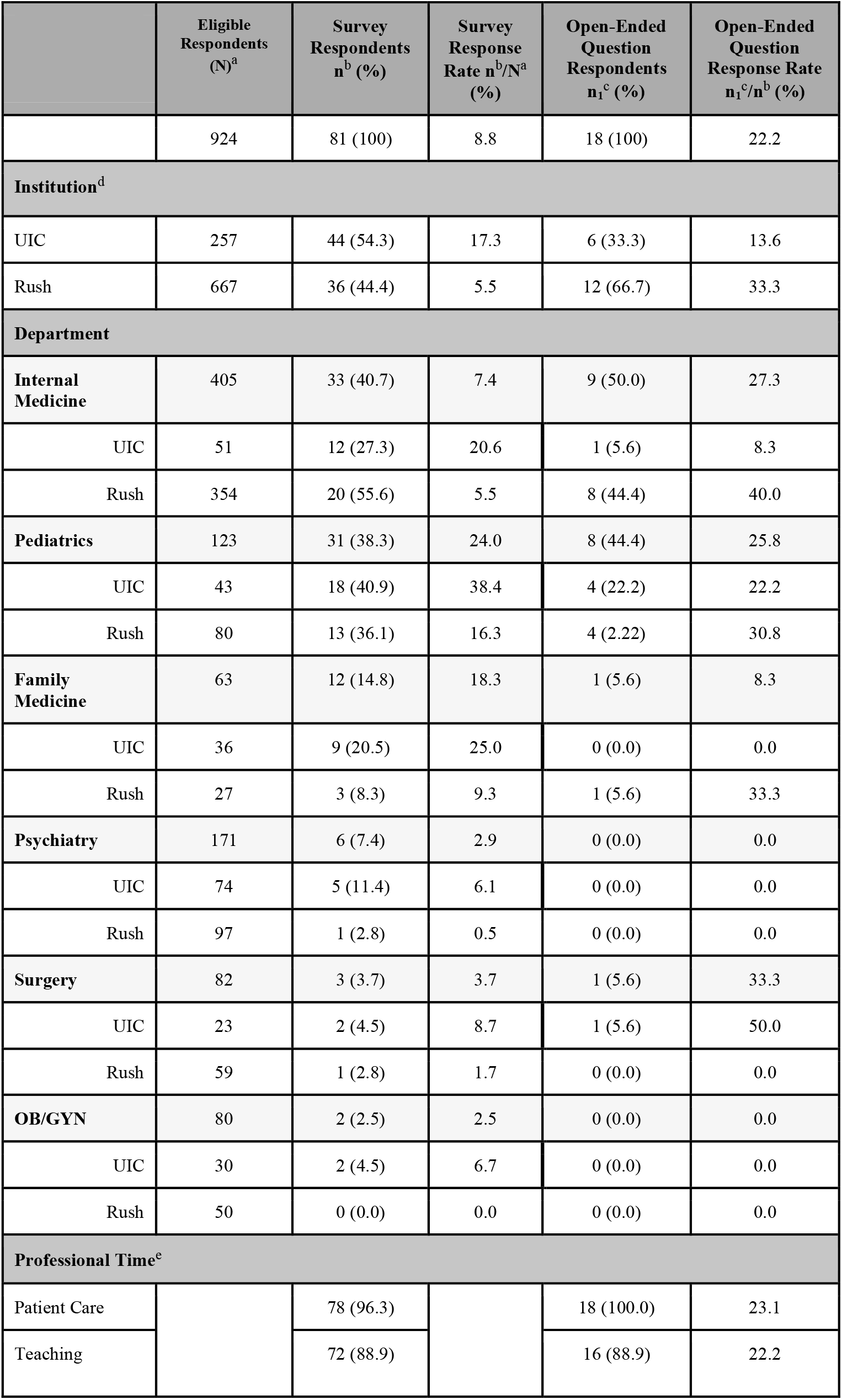
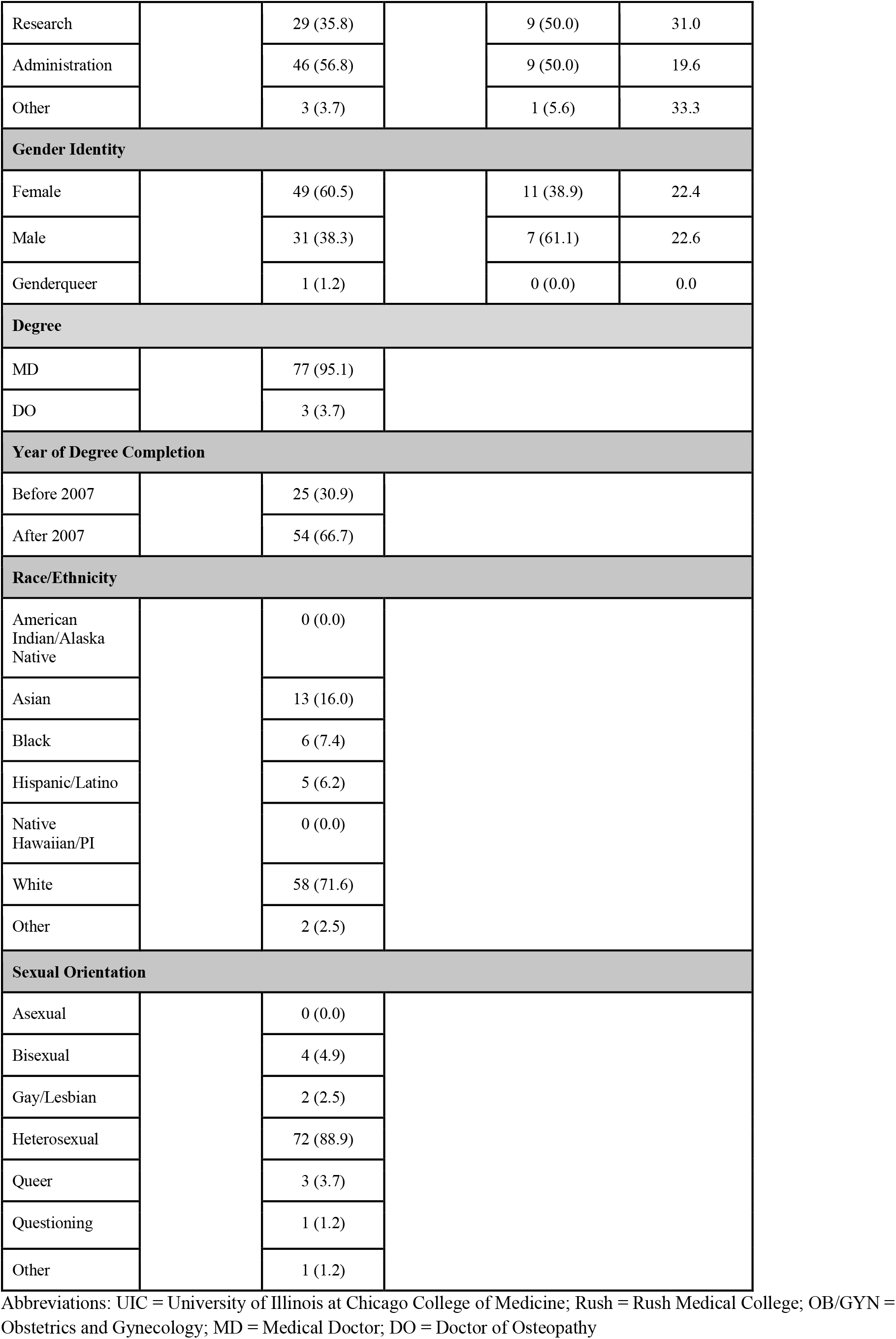
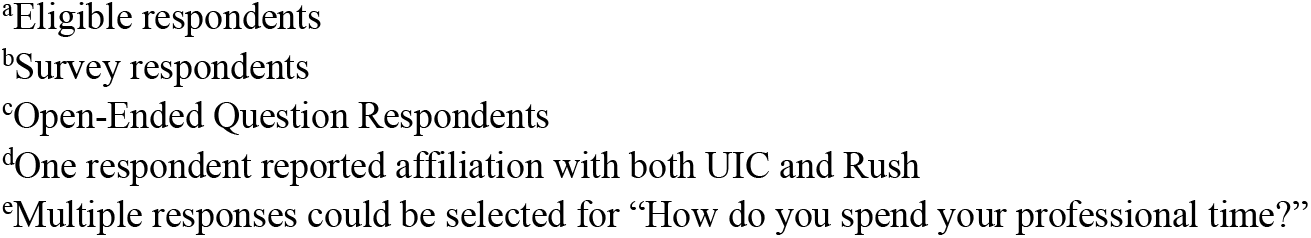
Response Rates and Demographics among 81 Medical Faculty Surveyed in 2020 in Chicago, IL

|  | Eligible Respondents (N)<sup>a</sup> | Survey Respondents n<sup>b</sup> (%) | Survey Response Rate n<sup>b</sup>/N<sup>a</sup> (%) | Open-Ended Question Respondents n<sup>c</sup> (%) | Open-Ended Question Response Rate n<sup>c</sup>/n<sup>b</sup> (%) |
| --- | --- | --- | --- | --- | --- |
|  | 924 | 81 (100) | 8.8 | 18 (100) | 22.2 |
| **Institution<sup>d</sup>** |  |  |  |  |  |
| UIC | 257 | 44 (54.3) | 17.3 | 6 (33.3) | 13.6 |
| Rush | 667 | 36 (44.4) | 5.5 | 12 (66.7) | 33.3 |
| **Department** |  |  |  |  |  |
| **Internal Medicine** | 405 | 33 (40.7) | 7.4 | 9 (50.0) | 27.3 |
| UIC | 51 | 12 (27.3) | 20.6 | 1 (5.6) | 8.3 |
| Rush | 354 | 20 (55.6) | 5.5 | 8 (44.4) | 40.0 |
| **Pediatrics** | 123 | 31 (38.3) | 24.0 | 8 (44.4) | 25.8 |
| UIC | 43 | 18 (40.9) | 38.4 | 4 (22.2) | 22.2 |
| Rush | 80 | 13 (36.1) | 16.3 | 4 (2.22) | 30.8 |
| **Family Medicine** | 63 | 12 (14.8) | 18.3 | 1 (5.6) | 8.3 |
| UIC | 36 | 9 (20.5) | 25.0 | 0 (0.0) | 0.0 |
| Rush | 27 | 3 (8.3) | 9.3 | 1 (5.6) | 33.3 |
| **Psychiatry** | 171 | 6 (7.4) | 2.9 | 0 (0.0) | 0.0 |
| UIC | 74 | 5 (11.4) | 6.1 | 0 (0.0) | 0.0 |
| Rush | 97 | 1 (2.8) | 0.5 | 0 (0.0) | 0.0 |
| **Surgery** | 82 | 3 (3.7) | 3.7 | 1 (5.6) | 33.3 |
| UIC | 23 | 2 (4.5) | 8.7 | 1 (5.6) | 50.0 |
| Rush | 59 | 1 (2.8) | 1.7 | 0 (0.0) | 0.0 |
| **OB/GYN** | 80 | 2 (2.5) | 2.5 | 0 (0.0) | 0.0 |
| UIC | 30 | 2 (4.5) | 6.7 | 0 (0.0) | 0.0 |
| Rush | 50 | 0 (0.0) | 0.0 | 0 (0.0) | 0.0 |
| **Professional Time<sup>e</sup>** |  |  |  |  |  |
| Patient Care |  | 78 (96.3) |  | 18 (100.0) | 23.1 |
| Teaching |  | 72 (88.9) |  | 16 (88.9) | 22.2 |
| Research |  | 29 (35.8) |  | 9 (50.0) | 31.0 |
| Administration |  | 46 (56.8) |  | 9 (50.0) | 19.6 |
| Other |  | 3 (3.7) |  | 1 (5.6) | 33.3 |
| **Gender Identity** |  |  |  |  |  |
| Female |  | 49 (60.5) |  | 11 (38.9) | 22.4 |
| Male |  | 31 (38.3) |  | 7 (61.1) | 22.6 |
| Genderqueer |  | 1 (1.2) |  | 0 (0.0) | 0.0 |
| **Degree** |  |  |  |  |  |
| MD |  | 77 (95.1) |  |  |  |
| DO |  | 3 (3.7) |  |  |  |
| **Year of Degree Completion** |  |  |  |  |  |
| Before 2007 |  | 25 (30.9) |  |  |  |
| After 2007 |  | 54 (66.7) |  |  |  |
| **Race/Ethnicity** |  |  |  |  |  |
| American Indian/Alaska Native |  | 0 (0.0) |  |  |  |
| Asian |  | 13 (16.0) |  |  |  |
| Black |  | 6 (7.4) |  |  |  |
| Hispanic/Latino |  | 5 (6.2) |  |  |  |
| Native Hawaiian/PI |  | 0 (0.0) |  |  |  |
| White |  | 58 (71.6) |  |  |  |
| Other |  | 2 (2.5) |  |  |  |
| **Sexual Orientation** |  |  |  |  |  |
| Asexual |  | 0 (0.0) |  |  |  |
| Bisexual |  | 4 (4.9) |  |  |  |
| Gay/Lesbian |  | 2 (2.5) |  |  |  |
| Heterosexual |  | 72 (88.9) |  |  |  |
| Queer |  | 3 (3.7) |  |  |  |
| Questioning |  | 1 (1.2) |  |  |  |
| Other |  | 1 (1.2) |  |  |  |

Abbreviations: UIC = University of Illinois at Chicago College of Medicine; Rush = Rush Medical College; OB/GYN = Obstetrics and Gynecology; MD = Medical Doctor; DO = Doctor of Osteopathy
<sup>a</sup>Eligible respondents
<sup>b</sup>Survey respondents
<sup>c</sup>Open-Ended Question Respondents
<sup>d</sup>One respondent reported affiliation with both UIC and Rush
<sup>e</sup>Multiple responses could be selected for “How do you spend your professional time?”

Table 2 summarizes the achievement of high scores in both knowledge and beliefs categories by the characteristics of the respondents. The average correct knowledge score was 78.0% (0.21). The average favorable beliefs score was 71.9% (0.18). Fifty-three (65.4%) respondents achieved high knowledge scores. Thirty-four (42.0%) respondents achieved high beliefs scores. Twenty-six (32.1%) achieved high scores in both categories.

**Table 2.** High Knowledge, Belief, and Concept Exposure Scores by Characteristics of 81 Medical Faculty Respondents Surveyed in 2020 in Chicago, IL

|  | Total<br>n<sup>a</sup> (%) | UIC<br>n<sup>a</sup> (%) | Rush<br>n<sup>a</sup> (%) | P-value | M<br>n<sup>a</sup> (%) | F<br>n<sup>a</sup> (%) | P-value | IM<br>n<sup>a</sup> (%) | Not<br>IM<br>n<sup>a</sup> (%) | P-value | Peds<br>n<sup>a</sup> (%) | Not<br>Peds<br>n<sup>a</sup> (%) | P-value | TC<br>Before<br>2007<br>n<sup>a</sup> (%) | TC<br>After<br>2007<br>n<sup>a</sup> (%) | P-value |
| --- | --- | --- | --- | --- | --- | --- | --- | --- | --- | --- | --- | --- | --- | --- | --- | --- |
| **Respondents** | 81<br>(100) | 44<br>(100) | 36<br>(100) |  | 31<br>(100) | 49<br>(100) |  | 33<br>(100) | 48<br>(100) |  | 31<br>(100) | 50<br>(100) |  | 25<br>(100) | 54<br>(100) |  |
| **High**<br>**Knowledge**<br>**Score** | 53<br>(65.4) | 30<br>(68.2) | 23<br>(63.9) | 0.35 | 15<br>(48.4) | 37<br>(75.5) | 0.01 | 20<br>(60.6) | 20<br>(60.6) | 0.45 | 24<br>(77.4) | 29<br>(58.0) | 0.07 | 17<br>(68.0) | 35<br>(64.8) | 0.78 |
| **High**<br>**Beliefs**<br>**Score** | 34<br>(42.0) | 22<br>(50.0) | 12<br>(33.3) | 0.22 | 10<br>(32.3) | 23<br>(46.9) | 0.19 | 10<br>(30.3) | 24<br>(50.0) | 0.08 | 15<br>(48.4) | 19<br>(38.0) | 0.36 | 9<br>(36.0) | 25<br>(46.3) | 0.39 |
| **High**<br>**Knowledge**<br>**+ Beliefs**<br>**Score** | 26<br>(32.1) | 16<br>(36.4) | 10<br>(27.8) | 0.56 | 7<br>(22.6) | 18<br>(36.7) | 0.18 | 8<br>(24.2) | 18<br>(37.5) | 0.21 | 14<br>(45.2) | 12<br>(24.0) | 0.05 | 8<br>(32.0) | 18<br>(33.3) | 0.91 |
| **High**<br>**Concept**<br>**Exposure**<br>**Score** | 42<br>(51.9) | 27<br>(61.4) | 15<br>(41.7) | 0.12 | 12<br>(38.7) | 29<br>(59.2) | 0.07 | 14<br>(42.4) | 28<br>(58.3) | 0.16 | 20<br>(64.5) | 22<br>(44.0) | 0.07 | 12<br>(48.0) | 29<br>(53.7) | 0.64 |

<sup>a</sup>Number of respondents
Abbreviations: UIC = University of Illinois at Chicago College of Medicine; Rush = Rush Medical College; M = Male; F = Female; IM = Internal Medicine; Peds = Pediatrics; TC = Training Completion

Respondents achieving high knowledge scores were significantly more likely to be female (37 respondents, 75.5%) than male (15 respondents, 48.4%) (P = .01). High knowledge scores did not differ significantly by respondent institution, specialty, or residency completion year. The most commonly missed items were “Childhood adversity is preventable”, which 42 (51.8%) of respondents missed, and “Patients who present with unexplained symptoms…often have a history of childhood adversity”, which 38 (46.9%) of respondents missed.

Although no respondent characteristics significantly correlated with attaining high beliefs scores, respondents who achieved both high knowledge and belief scores were significantly more likely to be pediatricians (14 respondents, 45.2%) compared to all other specialties (12 respondents, 24.0%) (P = .047). Respondents were least likely to answer the beliefs questions “I have direct access to resources that can help my patients prevent and/or address the consequences of childhood adversity”, which 55 (68.7%) respondents answered unfavorably and, “I don’t have time to ask patients about their childhood history”, which 49 (60.5%) answered unfavorably. See Appendix Table A for detailed survey answers.

Seventy-three respondents (90.1%) reported exposure to at least 6 (55.0%) of concepts, and 42 (51.9%) of respondents reported exposure to at least 80% (9 or more) concepts in some way (Table 2, Appendix Table B). Respondent institution, gender, specialty, or year of residency completion did not significantly affect exposure scores. Overall, respondents were less likely to report exposure to study concepts through their formal medical education than other routes. Seventeen respondents (21.0%) communicated that they had no formal exposure to any of the concepts, and only six (7.4%) selected formal exposure to at least 80% of concepts.

Through exposure of any kind, 78 (96.3%) of the respondents were familiar with the concepts, “Childhood experiences shape the growth, development, and structure of the brain”; “Individual, family, community, structural and historical childhood experiences…are powerful influences of health across the lifespan”; and “Human brains have the capacity for neuroplasticity…with the greatest capacity early in life.” In contrast, only 11 (13.6%) reported exposure to the concept that healthcare professionals are more likely to have experienced childhood adversity themselves.

Table 3 includes responses to the question, “Please indicate the relationship of the above concepts to your teaching, practice, research, and/or administrative work.” Although 78 (96.3%) of the respondents indicated that survey concepts are relevant to their work, 48 (59.3%) reported needing more coaching for full incorporation of these concepts, and only 18 (22.2%) reported fully incorporating them. Respondents reporting full incorporation were significantly more likely to have high concept exposure scores (17 respondents, 94.4%) compared with those not fully incorporating the concepts (25 respondents, 39.7 %) (P < .001), regardless of whether they reported formal or informal exposure. Respondents reporting that survey concepts are “relevant to my work, but I am not applying them” (mean agreement 4.1 (0.52)) agreed more strongly with the statement, “I don’t have time to ask patients about their childhood history” than respondents who stated the concepts were relevant, and they were applying them at least to some degree (mean agreement 2.7 (1.1)) (P = .001). Responses to the relevance question did not differ significantly in relation to agreement with the statement “I feel uncomfortable asking patients about their childhood history.”

**Table 3.**
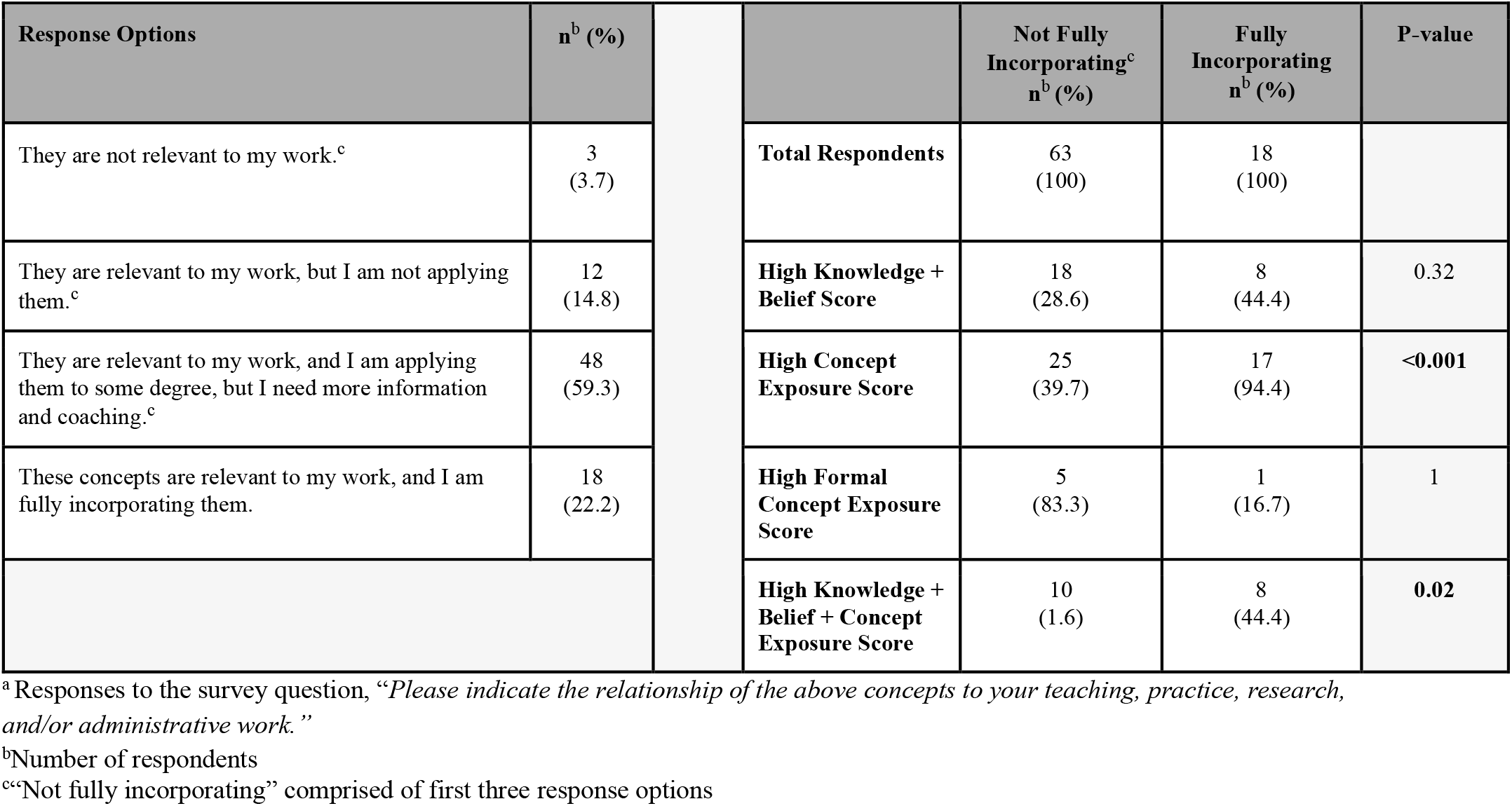
Perception of Relevance and Degree of Application^a^ vs. High Knowledge, Belief, and Concept Exposure Score Among 81 Medical Faculty Respondents Surveyed in 2020 in Chicago, IL

Of the 81 survey participants included in the data analysis, 21 (25.9%) responded to the optional open-ended question, “Please provide any additional comments, questions, or clarifications you would like to make about any of the material in the survey below”. See Table 1 for demographics of those who responded to the open-ended question. Respondents identifying themselves as internists, pediatricians, surgeons, men, Rush faculty, teachers, and researchers, as well as those describing the survey concepts as either not relevant to their practice or relevant to their practice but not being applied, had greater representation among respondents to the open-ended question than they had among full survey respondents. Of the 21 responses, our team excluded three (14.2%) from the qualitative analysis due to identifying information or unrelated subject matter. We categorized the remaining 18 (85.7%) into five overarching themes and a total of 10 sub-themes: Reflections and/or recommendations about professional life, including comments about patients, practice, understanding and application of evidence and specific specialties or conditions; Personal vignettes; Reflections and/or recommendations about relevant education to date as well as aspirations for the future; Reflections and/or recommendations about the survey itself; and Appreciation of the study topic area in general. Each response yielded between one and six themes/sub-themes. Professional Life, particularly Practice Challenges, and those regarding the survey itself were the most common themes/subthemes. Although some respondents commented about their own education around survey topics and/or expressed a desire/need to learn more, none commented about their role or experiences teaching these concepts. See Table 4 for sample comments, distribution of individual respondents by theme, and frequency of themes and subthemes.

**Table 4.**
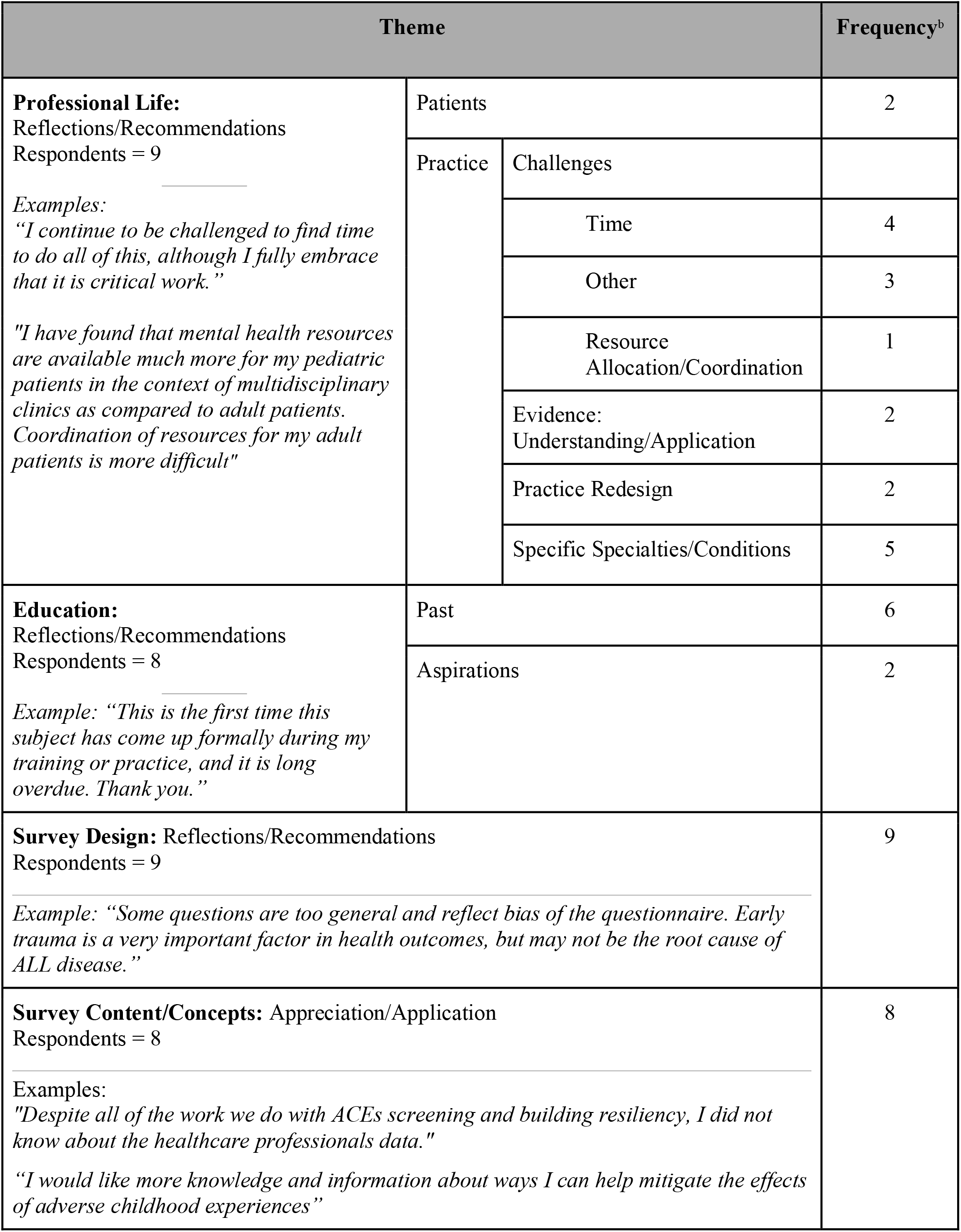
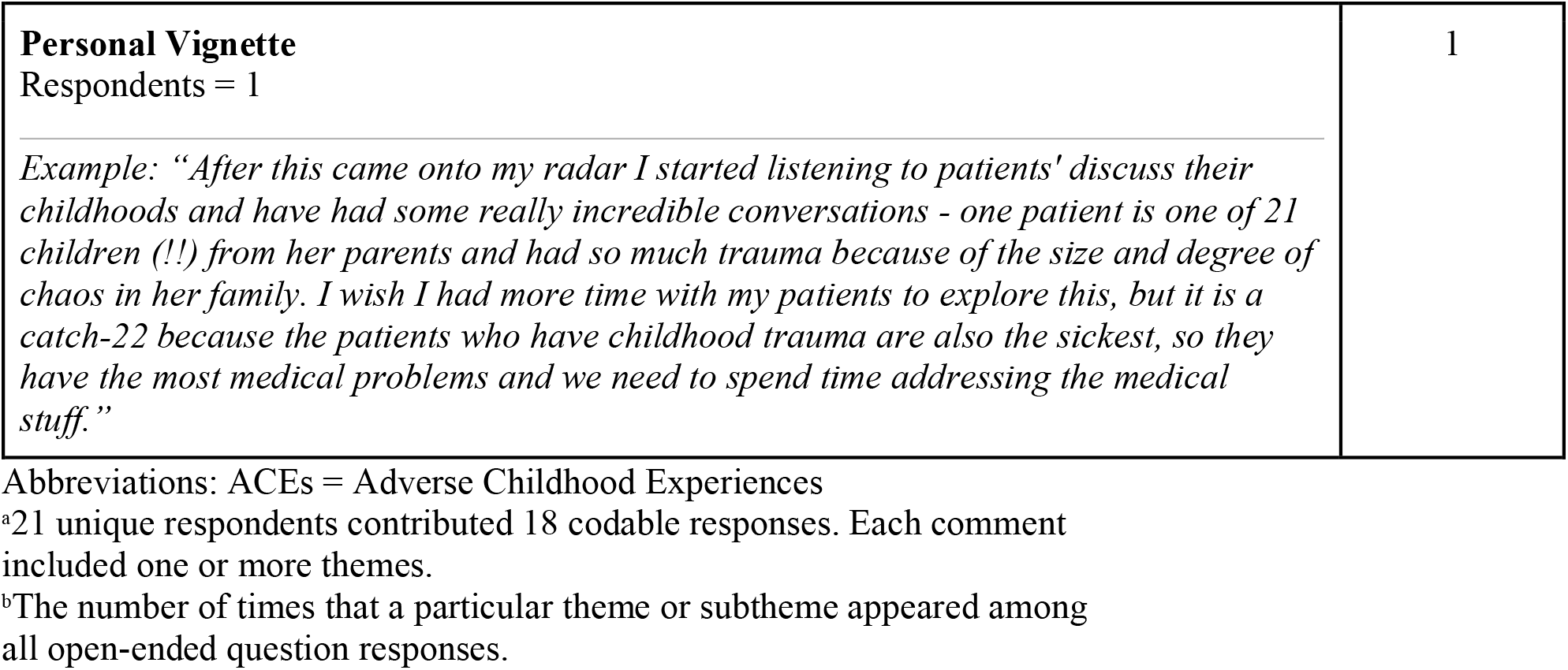
Themes and Subthemes Arising from Responses to an Optional, Open-Ended Question Included in a Survey of Medical Faculty^a^ in 2020 in Chicago, IL

## Discussion

To the best of our knowledge, this exploratory survey of six departments at two medical schools is the first to investigate physician faculty preparation to teach about the embodiment of childhood experiences and its influence on health across the life course. Our findings simultaneously show a moderate degree of knowledge among faculty about the impact of childhood experiences on health and suggest that additional training is necessary. In spite of the reality that few medical schools include any trauma education,^24^ more than half of survey respondents achieved high knowledge and concept exposure scores via various formal and informal routes. Almost all respondents reported that study concepts were relevant to their work, and many expressed incorporating them to some extent. Nonetheless, few reported fully incorporating study concepts and skills, and many identified the need for additional education and training, both in their responses to the relevance/application question and to the open-ended question. Of note, respondents who described full incorporation were also significantly more likely to achieve high concept exposure scores, emphasizing that faculty application of the science of trauma and healing appears to depend at least in part on exposure. High knowledge and beliefs scores seem to play a less significant role in full incorporation of concepts. Although surprising, this is perhaps reassuring--simply educating faculty, rather than attempting to change beliefs, may make the biggest difference in promoting practice change. As expected,^34-36^believing that one does not have enough time to inquire about patients’ childhood history significantly decreases the likelihood of fully applying concepts. Discomfort with inquiring about childhood experiences, however, while present for some respondents, was not an associated barrier.

Consistent with the dearth of relevant content in medical education,^24^ our study population reported that most of their exposure to study concepts and skills occurred outside formal training, yet the route of exposure did not affect respondents’ perception of relevance or their degree of applying the science connecting childhood experiences to health across the life course. Furthermore, respondents’ institution, gender, specialty, or year of residency completion were not significantly associated with exposure scores. Among those demographic differences that we were able to measure with confidence, few were significantly associated with high knowledge or belief score attainment, suggesting that all faculty can learn and apply this material.

Survey responses to both the multiple choice and open-ended portions of the study revealed specific gaps in faculty preparation regarding the preventable nature and clinical presentations of childhood adversity, prevalence and features of health care workers’ own trauma, effective evidence-based prevention and treatment strategies, and coping with multi-layered concerns of time constraints and insufficient resources. In responses to the open-ended question, participants reported considering or applying study concepts in their role as clinicians, but none reflected on their role in teaching this science to medical trainees. When medical educators themselves have knowledge gaps or do not see themselves as teaching particular content, both the quality of education and learner wellness suffer. Inadequate knowledge about the profound impact of childhood experiences on health and how to apply it diminishes the care faculty provide to their patients, their relationships with students and peers, and even their own well-being.

### Study limitations

Multiple factors may limit the generalizability of this study to the community of US medical faculty as a whole. Although our survey tool was based on peer-reviewed literature and developed through an iterative process utilizing cognitive interviews, no standard for assessing faculty preparation to teach survey concepts exists. Therefore, our survey’s ability to fully measure faculty preparation is still unexplored. Second, a small convenience sample, variation in response rates by institution and department, and low overall response rate limit the generalizability of the results. This survey served as an initial exploration, so limiting sample size and geography was intentional. The notable difference in eligible participants and response rates between UIH-UIC COM and Rush may result from variation in terms of which specialties and subspecialties appear in each institution’s department directories. UIC COM’s directory almost exclusively included generalists from the six eligible departments while Rush’s directory included specialists in addition to generalists.

Because survey participation was voluntary, physicians with interest in and/or familiarity with the survey topic may have been more likely to participate, hence contributing to self-selection bias. Interestingly, this was not the case for Psychiatry faculty in spite of their inevitable exposure to survey content in their daily work. Faculty from Pediatrics, Family Medicine, and Internal Medicine had the highest rates of survey participation, perhaps reflecting our team’s historical contact with and/or reputation among these departments. Thus, our study sample comprised of predominantly white, female, heterosexual faculty from Pediatrics, Family Medicine, and Internal Medicine may not represent all faculty either from departments eligible for this study or nationally.

Finally, multiple circumstances affecting survey dissemination likely contributed to the low overall participation rate. Restrictions to in-person gatherings in response to the Coronavirus (COVID-19) pandemic required us to deliver the survey solely via email, forcing us to miss planned collection opportunities at in-person department meetings. Our data collection period also coincided with a time of both predictable and unprecedented stress for medical faculty: predictable annual new resident onboarding as well as the unprecedented experiences of a prolonged global pandemic and the murder in late May 2020 of George Floyd, one among many Black Americans who face the consequences of centuries of racial injustice and structural violence, which has now become increasingly visible. All of these circumstances may have decreased survey participation rates.

### Future directions

Expanding the reach of this survey to include a representative national sample of medical schools and faculty to assess preparation to teach essential neurobiology, epidemiology, developmental psychology and traumatology concepts and skills, as well as to discern any difference in exposure to, and mastery of, survey concepts by institution, department, geographic region or other factors, will help better identify gaps and best practices. Each is a necessary piece of translating the relevant science into medical education and practice. To begin to build an evidence base, we prioritized surveying medical faculty due to their key role in directing the prevention, diagnosis and treatment of disease and in educating future physicians. Extending survey dissemination to other health professionals, however, is another worthwhile step toward creating trauma-informed healing-centered health care environments.

In both the multiple choice and open-ended portions of the survey, few respondents reported either receiving formal education on a majority of study concepts or having the capacity to fully integrate them into their work. Consequently, intentional faculty development to fill identified gaps is pivotal to ensure faculty preparation to teach. Among other topics, targeted education includes lessons on the physiologic embedding of childhood experiences and how it shapes the health trajectory;^4,5,6,11,12,45^ implementation of universal trauma precautions and sensitive open-ended trauma inquiry;^46-48^ utilization of reflective and self-regulation practices;^49-52^ and concrete institutional and community partnership development strategies to meet patients’ multidimensional needs.^53,54^ This kind of professional education can empower faculty to successfully teach survey concepts to trainees, ensuring that they can fully incorporate these concepts when they move into faculty and practicing physician roles.

Although exposure to the science and skills underlying trauma-informed healing-centered care outside formal training is valuable, creation and dissemination of core curricular competencies to guide formal training will help assure optimal and standardized understanding and application of these concepts by physicians and future faculty. Such competencies would focus on the embodiment of childhood experiences; the cultivation of protective factors; and prevention and treatment of childhood adversity. Curricular transformations in nursing and medicine have already begun. An expert panel designed and disseminated 88 evidence-informed trauma and resilience curricular competencies for undergraduate, graduate and mental health specialty nursing training.^55^ Trauma-Informed Healthcare Education and Research, a national interprofessional multidisciplinary group, has created competencies for undergraduate medical education and is planning validation.^56^ Although not the focus of this study, systemic changes must accompany recommended faculty and trainee education in order to reap their full benefit. These suggested changes include scribes and team-based care;^57^ enhanced access to mental health, social, and other wraparound supports;^58,59^ and implementation of a new health care paradigm focused on health rather than disease^60^ and built on person-centered, strengths-oriented, trauma-informed, healing-centered, socially just principles and practices.^61,62^

## Conclusions

Knowledge and application of the science underlying the profound impact of childhood experiences on health and healing across the life course are foundational ingredients of high-quality medical training and clinical care which are not yet present throughout medical education. Most faculty in this study value this science, acknowledge its relevance, incorporate it to some degree in their work and identify a need for more instruction. Investment in intentional professional development that supports faculty confidence and capacity to teach this essential material is one critical step toward ensuring that it will be universally present in the core medical curriculum and ultimately in teaching, practice and research.

## Data Availability

The authors confirm that the data supporting the findings of this study are available within the article and its supplementary materials.

## Acknowledgments

The authors wish to thank Dr. Alisa Seo-Lee, Dr. Salman Khan, and Dr. Saad Alvi for their participation in the recruitment and execution of the cognitive interview process. We would also like to thank all participating faculty.

## Disclosures

### Ethical approval

This study was approved by the institutional review board of the University of Illinois at Chicago and Rush University Medical Center.

### Disclaimers

None.

### Previous presentations

The abstract of an earlier version of this article was presented as part of a poster session at the virtual Integrative Medicine for the Underserved (IM4US) 2020 Conference, the virtual University of Illinois College of Medicine Research Forum 2020, and the virtual National Collaborative for Education to Address the Social Determinants of Health 2021 Annual Conference.

## Appendix

**Appendix Table A**. Responses to Knowledge and Beliefs Questions Among 81 Medical Faculty Surveyed in 2020 in Chicago, IL

**Appendix Table B**. Responses to Exposure Questions Among 81 Medical Faculty Surveyed in 2020 in Chicago, IL

## Section 1: Demographics

Questions 1-9: Age, Gender, Race, Sexuality, Department, Year of training completion, Degree accreditation: MD or DO, Professional time distribution, Institutional affiliation

## Section 2: Knowledge and Belief Questions

(answered using a 5-point Likert scale)

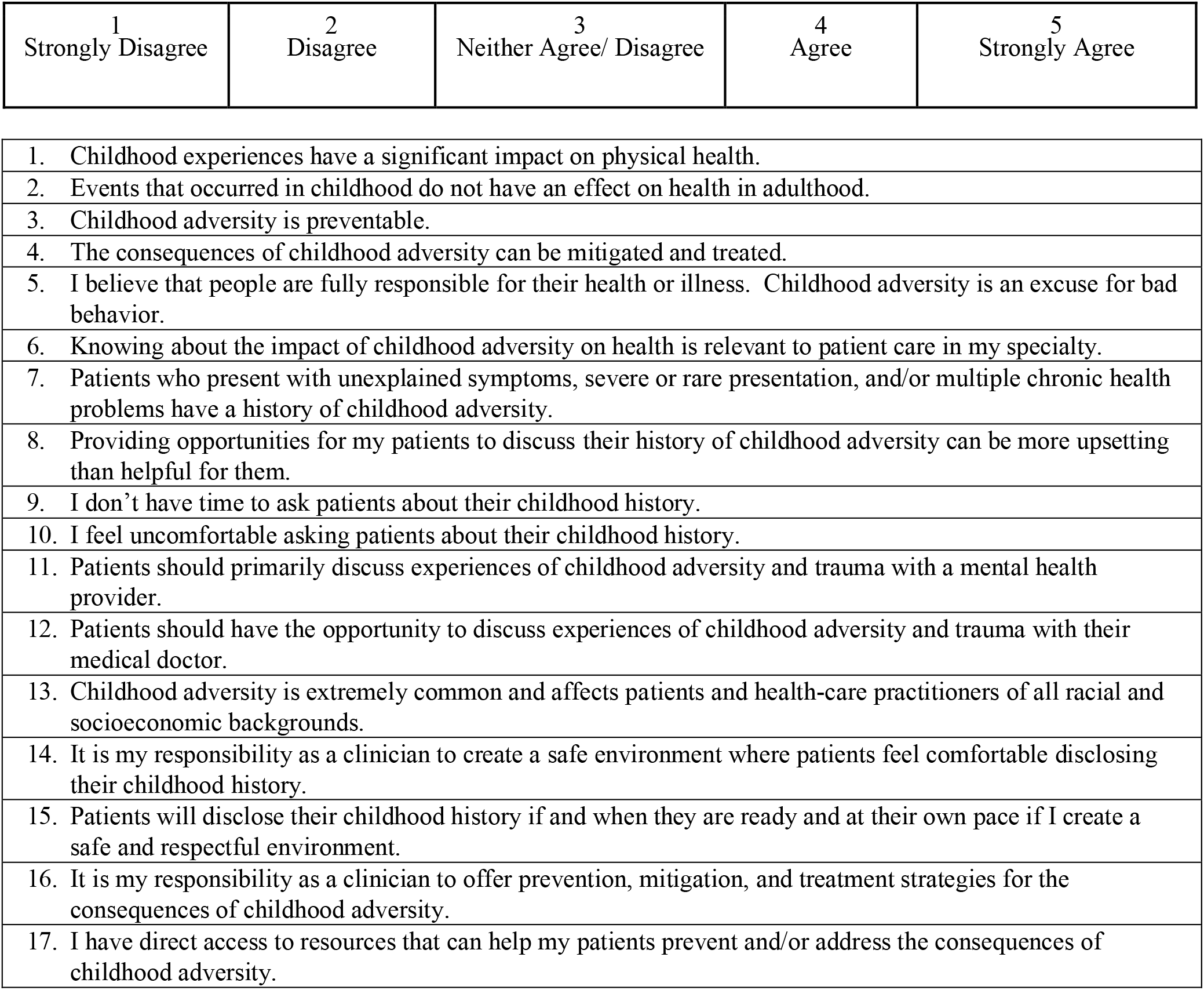

Section 2: Knowledge and Belief Questions (answered using a 5-point Likert scale)
| 1<br>Strongly Disagree | 2<br>Disagree | 3<br>Neither Agree/ Disagree | 4<br>Agree | 5<br>Strongly Agree |
| --- | --- | --- | --- | --- |

|  |
| --- |
| 1. Childhood experiences have a significant impact on physical health. |
| 2. Events that occurred in childhood do not have an effect on health in adulthood. |
| 3. Childhood adversity is preventable. |
| 4. The consequences of childhood adversity can be mitigated and treated. |
| 5. I believe that people are fully responsible for their health or illness. Childhood adversity is an excuse for bad behavior. |
| 6. Knowing about the impact of childhood adversity on health is relevant to patient care in my specialty. |
| 7. Patients who present with unexplained symptoms, severe or rare presentation, and/or multiple chronic health problems have a history of childhood adversity. |
| 8. Providing opportunities for my patients to discuss their history of childhood adversity can be more upsetting than helpful for them. |
| 9. I don't have time to ask patients about their childhood history. |
| 10. I feel uncomfortable asking patients about their childhood history. |
| 11. Patients should primarily discuss experiences of childhood adversity and trauma with a mental health provider. |
| 12. Patients should have the opportunity to discuss experiences of childhood adversity and trauma with their medical doctor. |
| 13. Childhood adversity is extremely common and affects patients and health-care practitioners of all racial and socioeconomic backgrounds. |
| 14. It is my responsibility as a clinician to create a safe environment where patients feel comfortable disclosing their childhood history. |
| 15. Patients will disclose their childhood history if and when they are ready and at their own pace if I create a safe and respectful environment. |
| 16. It is my responsibility as a clinician to offer prevention, mitigation, and treatment strategies for the consequences of childhood adversity. |
| 17. I have direct access to resources that can help my patients prevent and/or address the consequences of childhood adversity. |

## Section 3: History/content of knowledge acquisition

(Choose **<u>all</u>** that apply for each of the following statements:)

❏ I have ***not been exposed*** to this concept.
❏ I ***have been exposed*** to this concept, but I don’t remember when or where I learned it
❏ I was exposed to this concept ***within my formal training*** by being taught in
  ❏ undergraduate course(s)
  ❏ medical school course(s) or clinical rotation(s)
  ❏ formal residency education
  ❏ formal fellowship education
❏ I learned about this concept ***outside of my formal training***
  ❏ through journal article(s)
  ❏ from colleagues
  ❏ at conference(s)
  ❏ through the popular media
  ❏ from family or friends
  ❏ from listening to and observing my patients

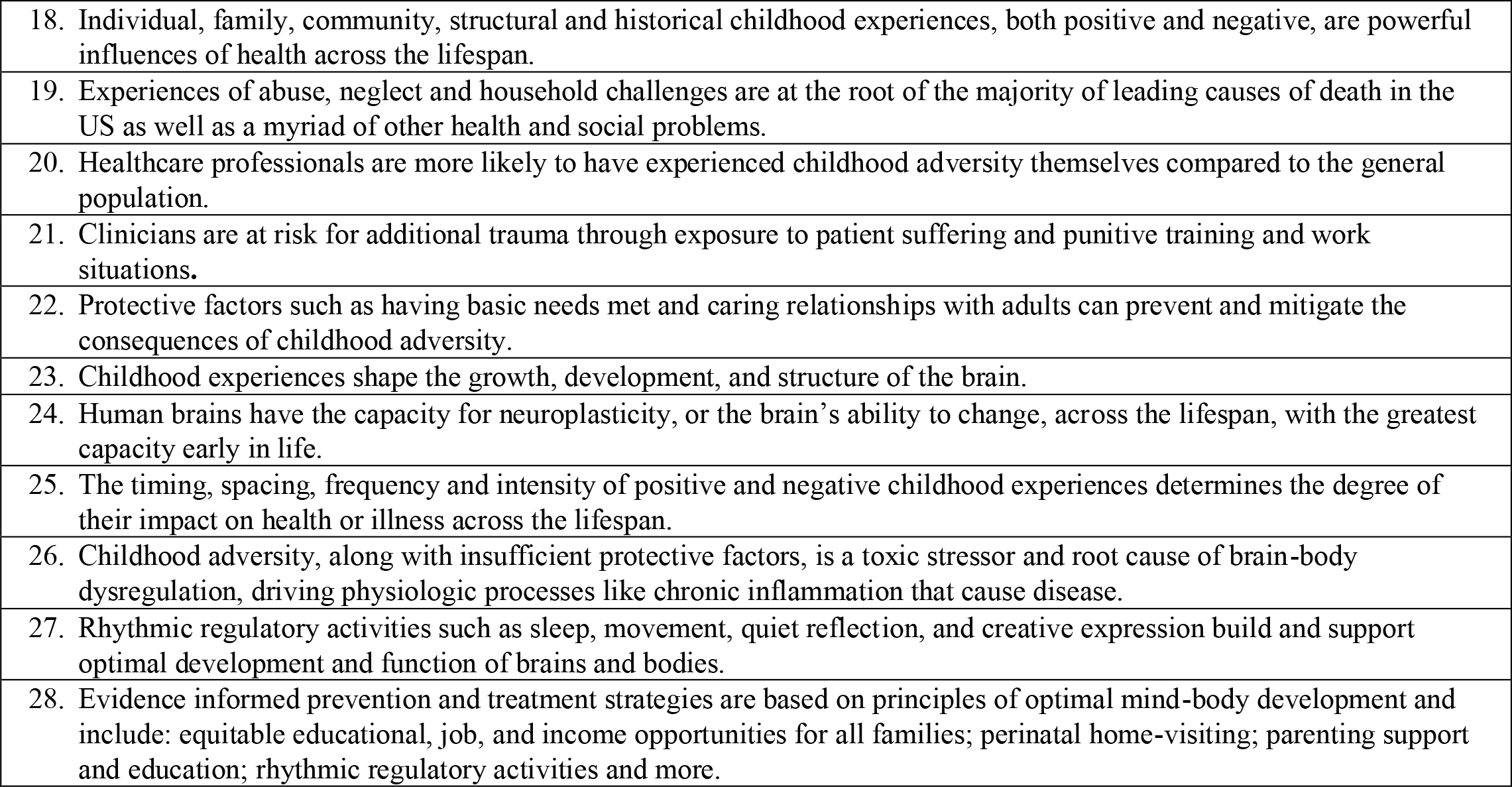

## Other/Optional

29. Please indicate the relationship of the above concepts to your teaching, practice, research, and/or administrative work.
  ❏ They are not relevant to my work
  ❏ They are relevant to my work, but I am not applying them.
  ❏ They are relevant to my work, and I am applying them to some degree, but I need more information and coaching.
  ❏ These concepts are relevant to my work, and I am fully incorporating them.
30. Please provide any additional comments, questions, or clarifications you would like to make about any of the material in the survey below.

