## Appendix Tables A and B for "Is academic medicine prepared to teach about the intersection of childhood experiences and health across the life course? An exploratory survey of faculty"

**Appendix Table A.** Responses to Knowledge and Beliefs Questions Among 81 Medical Faculty Surveyed in 2020 in Chicago, IL

| **Statement** | **K or B** | **1 Strongly Disagree n[a] (%)** | **2 Disagree n[a] (%)** | **3 Neither Agree nor Disagree n[a] (%)** | **4 Agree n[a] (%)** | **5 Strongly Agree n[a] (%)** | **Mean (SD)** | **"Correct"/ "Favorable" Response Rate (%)** |
|---|---|---|---|---|---|---|---|---|
| 1. Childhood experiences have a significant impact on physical health. | K | 0/81 (0.0) | 0/81 (0.0) | 4/81 (4.9) | 27/81 (33.3) | 50/81 (61.7) | 4.57 (0.59) | **95.1** |
| 2. Events that occurred in childhood do not have an effect on health in adulthood. | K | 59/81 (72.8) | 17/81 (21.0) | 2/81 (2.5) | 1/81 (1.2) | 2/81 (2.5) | 1.40 (0.82) | **93.8** |
| 3. Childhood adversity is preventable. | K | 1/81 (1.2) | 14/81 (17.3) | 27/81 (33.3) | 36/81 (44.4) | 3/81 (3.7) | 3.32 (0.85) | **48.2** |
| 4. The consequences of childhood adversity can be mitigated and treated. | K | 0/81 (0.0) | 1/81 (1.2) | 9/81 (11.1) | 60/81 (74.1) | 11/81 (13.6) | 4.00 (0.55) | **87.7** |
| 5. I believe that people are fully responsible for their health or illness. Childhood adversity is an excuse for bad behavior. | B | 48/81 (59.3) | 27/81 (33.3) | 5/81 (6.2) | 1/81 (1.2) | 0/81 (0.0) | 1.49 (0.67) | **92.6** |
| 6. Knowing about the impact of childhood adversity on health is relevant to patient care in my specialty. | B | 1/81 (1.2) | 0/81 (0.0) | 8/81 (9.9) | 23/81 (28.4) | 49/81 (60.5) | 4.47 (0.78) | **88.9** |
| 7. Patients who present with unexplained symptoms, severe or rare presentations, and/or multiple chronic health problems often have a history of childhood adversity. | K | 1/81 (1.2) | 1/81 (1.2) | 36/81 (44.4) | 31/81 (38.3) | 12/81 (14.8) | 3.64 (0.80) | **53.1** |

| | | | | | | | | |
|---|---|---|---|---|---|---|---|---|
| 8. Providing opportunities for my patients to discuss their history of childhood adversity can be more upsetting than helpful for them. | B | 16/81 (19.8) | 40/81 (49.4) | 23/81 (28.4) | 2/81 (2.5) | 0/81 (0.0) | 2.14 (0.75) | **69.1** |
| 9. I don't have time to ask patients about their childhood history. | B | 12/81 (14.8) | 20/81 (24.7) | 16/81 (19.8) | 30/81 (37.0) | 3/81 (3.7) | 2.90 (1.17) | **39.5** |
| 10. I feel uncomfortable asking patients about their childhood history. | B | 18/81 (22.2) | 38/81 (46.9) | 14/81 (17.3) | 10/81 (12.3) | 1/81 (1.2) | 2.23 (0.98) | **69.1** |
| 11. Patients should primarily discuss experiences of childhood adversity and trauma with a mental health provider. | B | 9/79 (11.4) | 35/79 (44.3) | 19/79 (24.1) | 15/79 (19.0) | 1/79 (1.3) | 2.54 (0.97) | **55.7** |
| 12. Patients should have the opportunity to discuss experiences of childhood adversity and trauma with their medical doctor. | B | 0/80 (0.0) | 1/80 (1.2) | 1/80 (1.2) | 38/80 (47.5) | 40/80 (50.0) | 4.46 (0.59) | **97.5** |
| 13. Childhood adversity is extremely common and affects patients and health-care practitioners of all racial and socioeconomic backgrounds. | K | 0/80 (0.0) | 0/80 (0.0) | 7/80 (8.8) | 30/80 (37.5) | 43/80 (53.8) | 4.45 (0.65) | **91.3** |
| 14. It is my responsibility as a clinician to create a safe environment where patients feel comfortable disclosing their childhood history. | B | 0/80 (0.0) | 0/80 (0.0) | 1/80 (1.2) | 24/80 (30.0) | 55/80 (68.8) | 4.67 (0.50) | **98.8** |
| 15. Patients will disclose their childhood history if and when they are ready and at their own pace if I create a safe and respectful environment. | B | 0/80 (0.0) | 1/80 (1.2) | 17/80 (21.2) | 36/80 (45.0) | 26/80 (32.5) | 4.09 (0.77) | **77.5** |

| | | | | | | | | |
|---|---|---|---|---|---|---|---|---|
| 16. It is my responsibility as a clinician to offer prevention, mitigation, and treatment strategies for the consequences of childhood adversity. | B | 1/79 (1.3) | 3/79 (3.8) | 13/79 (16.5) | 34/79 (43.0) | 28/79 (35.4) | 4.08 (0.89) | **78.5** |
| 17. I have direct access to resources that can help my patients prevent and/or address the consequences of childhood adversity. | B | 9/80 (11.2) | 25/80 (31.2) | 21/80 (26.2) | 17/80 (21.2) | 8/80 (10.0) | 2.88 (1.17) | **31.3** |

Abbreviations: K = Knowledge; B = Belief; SD = Standard Deviation

[a]Number of respondents

**Appendix Table B.** Responses to Exposure Questions Among 81 Medical Faculty Surveyed in 2020 in Chicago, IL

| Statement | Exposure Score[a] (%) | | n[b]/N[c] (%) | | n[b]/N[c] (%) |
|---|---|---|---|---|---|
| 18. Individual, family, community, structural and historical childhood experiences, both positive and negative, are powerful influences of health across the lifespan. | **96.3** | Formal Training | 33/86 (38.4) | Informal training | 35/86 (40.7) |
| | | Undergrad | 7 | Not sure where exposed | 15/86 (17.4) |
| | | Med School | 21 | No Exposure | 3/86 (3.5) |
| | | Residency | 30 | | |
| | | Fellowship | 9 | | |
| 19. Experiences of abuse, neglect and household challenges are at the root of the majority of leading causes of death in the US as well as a myriad of other health and social problems. | **77.8** | Formal Training | 26/87 (29.9) | Informal training | 26/87 (29.9) |
| | | Undergrad | 3 | Not sure where exposed | 17/87 (19.5) |
| | | Med School | 8 | No Exposure | 18/87 (20.7) |
| | | Residency | 24 | | |
| | | Fellowship | 8 | | |
| 20. Healthcare professionals are more likely to have experienced childhood adversity themselves compared to the general population. | **13.6** | Formal Training | 3/78 (3.8) | Informal training | 1/78 (1.3) |
| | | Undergrad | 1 | Not sure where exposed | 4/78 (5.1) |
| | | Med School | 1 | No Exposure | 70/78 (89.7) |
| | | Residency | 3 | | |
| | | Fellowship | 1 | | |
| 21. Clinicians are at risk for additional trauma through exposure to patient suffering and punitive training and work situations. | **71.6** | Formal Training | 12/79 (15.2) | Informal training | 17/79 (21.5) |
| | | Undergrad | 1 | Not sure where exposed | 27/79 (34.2) |
| | | Med School | 5 | No Exposure | 23/79 (29.1) |
| | | Residency | 10 | | |
| | | Fellowship | 3 | | |

| | | | | | |
|---|---|---|---|---|---|
| 22. Protective factors such as having basic needs met and caring relationships with adults can prevent and mitigate the consequences of childhood adversity. | **90.1** | Formal Training<br>Undergrad<br>Med School<br>Residency<br>Fellowship | 23/84 (27.4)<br>6<br>15<br>17<br>6 | Informal training<br>Not sure where exposed<br>No Exposure | 28/84 (33.3)<br>25/84 (29.8)<br>8/84 (9.5) |
| 23. Childhood experiences shape the growth, development, and structure of the brain. | **96.3** | Formal Training<br>Undergrad<br>Med School<br>Residency<br>Fellowship | 39/84 (46.4)<br>8<br>24<br>24<br>8 | Informal training<br>Not sure where exposed<br>No Exposure | 25/84 (29.8)<br>17/84 (20.2)<br>3/84 (3.6) |
| 24. Human brains have the capacity for neuroplasticity, or the brain's ability to change, across the lifespan, with the greatest capacity early in life. | **96.3** | Formal Training<br>Undergrad<br>Med School<br>Residency<br>Fellowship | 56/83 (67.5)<br>17<br>45<br>29<br>14 | Informal training<br>Not sure where exposed<br>No Exposure | 15/83 (18.1)<br>9/83 (10.8)<br>3/83 (3.6) |
| 25. The timing, spacing, frequency and intensity of positive and negative childhood experiences determines the degree of their impact on health or illness across the lifespan. | **69.1** | Formal Training<br>Undergrad<br>Med School<br>Residency<br>Fellowship | 17/80 (21.3)<br>3<br>7<br>11<br>4 | Informal training<br>Not sure where exposed<br>No Exposure | 19/80 (23.8)<br>19/80 (23.8)<br>25/80 (31.3) |
| 26. Childhood adversity, along with insufficient protective factors, is a toxic stressor and root cause of brain-body | **69.1** | Formal Training<br>Undergrad | 19/83 (22.9)<br>2 | Informal training<br>Not sure where exposed | 26/83 (31.3)<br>13/83 (15.7) |

| | | | | | |
|---|---|---|---|---|---|
| dysregulation, driving physiologic processes like chronic inflammation that cause disease. | | Med School | 5 | No Exposure | 25/83 (30.1) |
| | | Residency | 15 | | |
| | | Fellowship | 4 | | |
| 27. Rhythmic regulatory activities such as sleep, movement, quiet reflection, and creative expression build and support optimal development and function of brains and bodies. | **85.2** | Formal Training | 30/81 (37.0) | Informal training | 19/81 (23.5) |
| | | Undergrad | 3 | Not sure where exposed | 20/81 (24.7) |
| | | Med School | 17 | No Exposure | 12/81 (14.8) |
| | | Residency | 24 | | |
| | | Fellowship | 9 | | |
| 28. Evidence informed prevention and treatment strategies are based on principles of optimal mind-body development and include: equitable educational, job, and income opportunities for all families; perinatal home-visiting; parenting support and education; rhythmic regulatory activities and more. | **65.4** | Formal Training | 18/81 (22.2) | Informal training | 19/81 (23.5) |
| | | Undergrad | 3 | Not sure where exposed | 16/81 (19.8) |
| | | Med School | 6 | No Exposure | 28/81 (34.6) |
| | | Residency | 13 | | |
| | | Fellowship | 3 | | |

Abbreviations: US = United States; Undergrad = Undergraduate; Med School = Medical School

[a]Percent of Respondents with any exposure to concept: 100 x [(Total Respondents - Respondents Selecting “No Exposure”)/81] where Total Respondents=81 Respondents who completed the survey.

[b]Number of responses per type of exposure

[c]Total number of responses
